# Predictors of malaria prevention practices among HIV-positive patients attending an antiretroviral therapy clinic in Kampala, Uganda: a cross-sectional study

**DOI:** 10.64898/2026.10.08.26365025

**Authors:** Ambrose Ayebazibwe, Moses Kabangi, Joseph Mwaka

## Abstract

**Background:** Malaria and HIV remain major causes of illness in sub-Saharan Africa, accounting for 94% of the global malaria burden and over two-thirds of the global HIV burden. HIV-related immunosuppression increases susceptibility to malaria, with a higher malaria burden of up to 47% and severe malaria complications compared to HIV-negative individuals. Socio-demographic factors, individual knowledge, attitude, and perception are recognized determinants of malaria prevention behaviours, but no study has modelled their combined contribution, and there is scant research on these factors among people living with HIV. We assessed the level of knowledge, attitudes, and perceptions towards malaria, and identified the predictors of malaria prevention practice among HIV-positive individuals in Kampala, Uganda.

**Methods:** This cross-sectional study enrolled 620 HIV-positive patients attending the ART clinic of China-Uganda Friendship Hospital, Naguru, selected by systematic random sampling. Data collection was conducted from 21st July 2025 to 18th August 2025 using a structured, interviewer-administered questionnaire that measured socio-demographic characteristics, knowledge, attitude, perception, and the practice of malaria prevention. Associations were assessed using Mann-Whitney U, Kruskal-Wallis H, and Spearman correlation. The independent predictors of good malaria prevention practice were identified using binary logistic regression.

**Results:** Participants had a mean age of 34.8 years, were predominantly female (68.9%), Baganda (38.1%) by tribe, and the majority had a secondary level of education(46.0%). Most had good knowledge of malaria (97.3%) and a positive attitude (90.5%), while 57.3% held a positive perception of malaria risk and prevention; only 62.6% reported good malaria prevention practice, and most prevention methods used were sleeping under a mosquito net (71.1 3%) and closing windows early in the evening (70.3%). In regression analysis, knowledge (adjusted odds ratio [AOR] 1.198, 95% CI 1.03-1.40, p = 0.023) and attitude (AOR 1.101, 95% CI 1.04-1.17, p = 0.001) independently predicted good practice, while perception did not (AOR 1.00, 95% CI 0.96-1.04, p = 0.994). Scores of Attitude, perception, and malaria prevention practices varied significantly, though non-linearly, by education level, and were highest among non-educated participants.

**Conclusions:** Despite near-universal knowledge and predominantly positive attitudes in this study, more than a third of HIV-positive participants did not practice adequate malaria prevention, and perception did not independently predict malaria prevention practice once knowledge and attitude were accounted for. Programmes seeking to close this know-do gap should pair health education with structural support for prevention tools rather than education alone.

**Trial registration:** Not applicable. This is an observational study of an existing care population, not a healthcare intervention.

## Background

Malaria and human immunodeficiency virus (HIV) remain two of the most persistent public health threats in sub-Saharan Africa, and their co-occurrence in the same geographical area and individual compounds the burden of both diseases(1). In 2023, malaria affected an estimated 263 million people worldwide, with Africa accounting for 94% of cases and 95% of deaths. Uganda ranked third globally for malaria case burden, with about 1.3 million cases accounting for 4.8% of this global burden(2). In the same period, out of the 39.9 million people living with HIV globally, Africa carried more than two-thirds of the global HIV burden, and Uganda’s national HIV prevalence stood at approximately 5% by the end of December 2024, equivalent to roughly 1.4 to 1.5 million people living with HIV and new infections at 3700 which translates to 711 new infections weekly or 101 infections daily(3–5).

HIV-related immunosuppression, driven by progressive depletion of CD4+ T lymphocytes and impaired function of innate effector cells such as natural killer cells, reduces the host’s capacity to contain Plasmodium infection(6–10). This interaction underlies the wide range of malaria-HIV co-infection prevalence reported across the region, from single digits up to 47.7% depending on setting and population(1,11), with a pooled estimate of 22.7% among HIV-positive adults in sub-Saharan Africa [8]. Persons living with HIV had higher morbidity, mortality, and severe complications of malaria compared to HIV Negative persons (11–15), thus, further stressing the need to understand the factors responsible.

Malaria prevention depends not only on the availability of tools and methods such as insecticide-treated nets, repellents, indoor residual spraying, chemoprevention, and malaria vaccines (16–20) but also on individual knowledge, attitude, and perception(21–23). Perception is broadly captured by the Health Belief Model(HBM), in which perception, with constructs of perceived susceptibility, severity, benefits, and barriers, together shape whether protective behaviour is adopted (24–26). In combination with HBM, the KAP theory, as it is widely used(27–29), enabled us to further assess how knowledge and attitude lead to the practice of malaria prevention behaviours. The evidence for how these three domains operate, singly and in combination, specifically among people living with HIV, remains thin. A Nigerian study of HIV-positive adults examined knowledge of net use alone(22); a related Nigerian study examined knowledge and perception but not attitude (21). To our knowledge, no published study has modelled the independent, combined contribution of knowledge, attitude, and perception to malaria prevention practice within a single HIV-positive cohort, and this question had not previously been examined among HIV-positive adults in Uganda.

Understanding which of these domains actually changes behaviour once modelled together, rather than in isolation, is what should determine where limited health education resources are targeted. We therefore assessed the level of knowledge, attitude, perception, and predictors of the practice of malaria prevention among HIV-positive adults attending the ART clinic of China-Uganda Friendship Hospital, Naguru, Kampala, and used multivariable modelling to identify which of these domains, together with socio-demographic characteristics, independently predict malaria prevention practice.

## Methods

### Study design, setting, and population

This was a cross-sectional, quantitative study conducted among HIV-positive patients receiving care at the ART clinic of China-Uganda Friendship Hospital (also known as Naguru Regional Referral Hospital), a 100-bed public referral facility in Nakawa Division, Kampala, Uganda, whose ART clinic serves approximately 100 patients per weekday. Data were collected between 21st July 2025 and 18th August 2025. Eligible participants were HIV-positive patients aged 8 years and older who provided written informed consent. Written assent and parental or guardian consent were obtained for participants under 18 years; patients who were unable to speak or too unwell were excluded.

### Sample size and sampling

The minimum sample size was calculated for a single proportion using a pooled malaria prevalence among HIV-positive adults in sub-Saharan Africa of 22.7% [8], 95% confidence, and 5% precision, giving 551 participants; inflating for an anticipated 10% non-response gave a target of 613. Participants were recruited by systematic random sampling from the clinic’s daily screening log, using an accessible population of approximately 2,200 patients per month and a sampling interval of 4, with a random start between 1 and 4. Number 2 was the starting random number obtained from a calculator, and every 4th person was picked to provide consent. A total of 620 participants were enrolled.

### Measures

Data were collected using a structured, interviewer-administered questionnaire, translated into Luganda and programmed in KoBoToolbox. The instrument had five sections: socio-demographic characteristics (8 items); knowledge of malaria (10 yes/no items, 1 point each, total range 0-10, categorised as poor [0-5] or good [6-10]); attitude toward malaria prevention (7 items on a 4-point Likert scale, reverse-scored for negatively worded statements, total range 0-21 points, categorised as negative Attitude [≤ 10.5] or positive Attitude [> 10.5]); perception of malaria risk and prevention, covering perceived susceptibility, severity, benefits, and barriers drawn from the Health Belief Model (20 items, 4-point Likert scale, total range 0-60 points, categorised as negative perception [≤ 30] or positive perception [> 30]); and malaria prevention practice (9 items on a 4-point likert scale, total range 0-27 points, categorised as poor [0-13.5] or good [13.6-27]). The instrument was developed from a review of published knowledge-attitude-practice and Health Belief Model instruments, pretested on 10% of the target sample, and each section had a Cronbach’s alpha above 0.7.

### Ethical considerations

Ethical approval was obtained from the Research Ethics Committee at Kabale University (protocol KABREC-2025-654), with additional clearance from the research committee of China-Uganda Friendship Hospital, Naguru. All participants gave written informed consent; participants under 18 years provided assent together with parental or guardian consent. Consent forms were stored separately from completed questionnaires to preserve anonymity, and all electronic data were held on a password-protected database accessible only to study staff.

### Statistical analysis

Data were exported from KoBoToolbox, cleaned in Excel, and analysed in SPSS version 21. Descriptive statistics summarised socio-demographic characteristics and the level of each knowledge, attitude, perception, and practice domain. Associations between sex and each domain were tested using the Mann-Whitney U test, and associations between education level and each domain were tested using the Kruskal-Wallis H test with Bonferroni-corrected pairwise comparisons. Associations among knowledge, attitude, perception, and practice scores were tested using Spearman’s rank correlation. Knowledge, attitude, and perception scores were entered simultaneously into a binary logistic regression model with malaria prevention practice (good versus poor practice) as the outcome; predictors were evaluated at alpha = 0.05, and model fit was assessed with the omnibus likelihood-ratio test and the Hosmer-Lemeshow goodness-of-fit test.

## Results

### Socio-demographic characteristics

Of 620 participants enrolled, the mean age was 34.8 years (range 14-85), and 56.5% were aged 20 to 39 years. Women made up 68.9% of the sample, and Baganda made up the majority ethnic group at 38.1%. Just under half (46.0%) had attained secondary education, 30.2% primary, 19.8% tertiary, and 4.0% no formal education. Most participants were single (42.7%) or married (39.7%), while business persons (31.0%) and agriculture or casual labour (22.4%) were the most common occupations (Table 1).

**Table 1.**
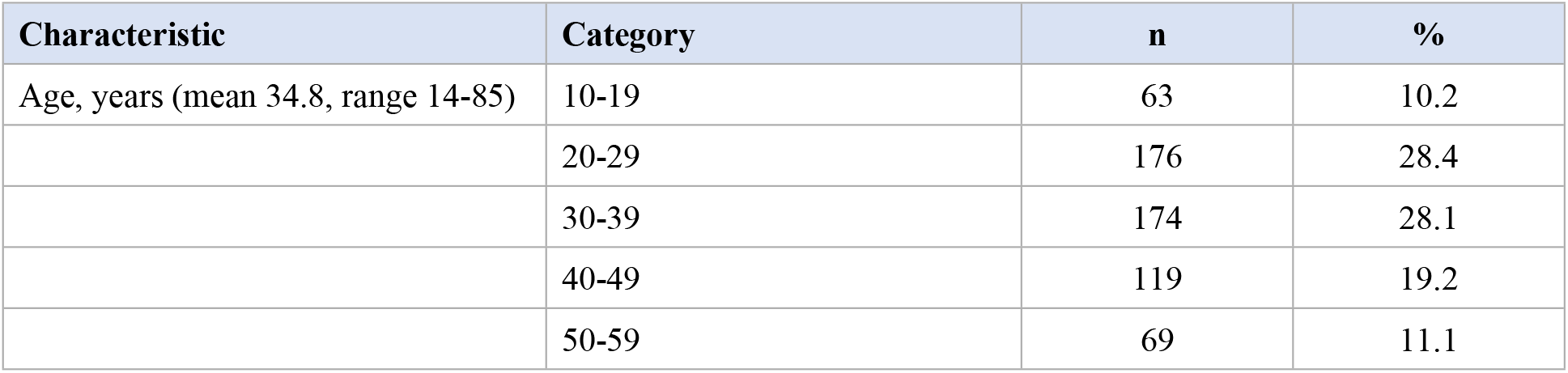
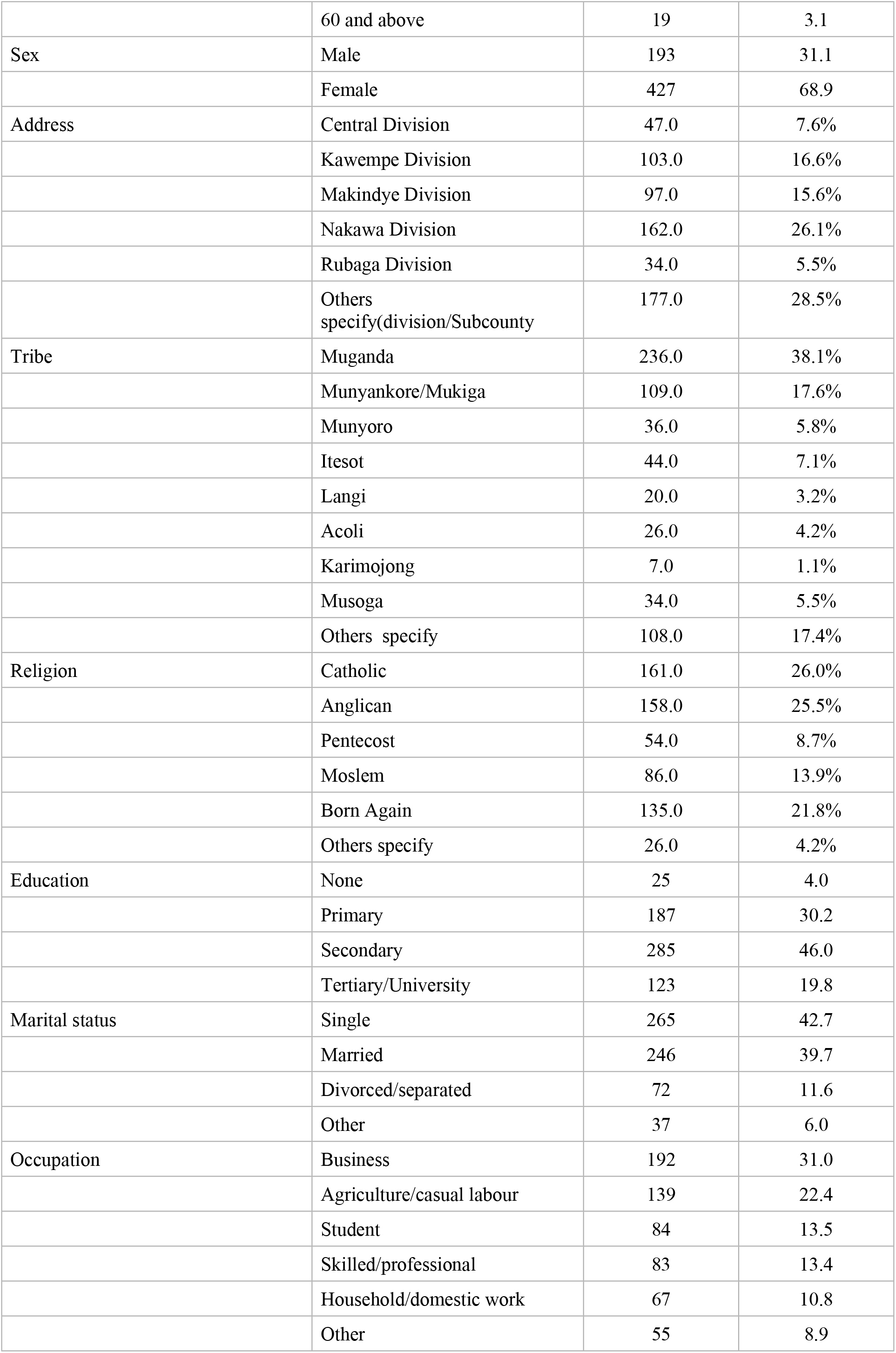
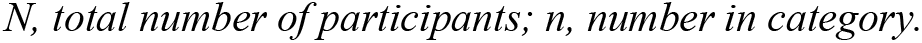
Socio-demographic characteristics of participants (N = 620)

### Level of knowledge, attitude, perception, and malaria prevention practice

Almost all participants had good knowledge of malaria (97.3%; mean score 8.8/10, SD 1.3) and a positive attitude toward malaria prevention (90.5%; mean 16.2/21, SD 3.7). A smaller majority, 57.3%, held a positive perception of malaria risk and prevention (mean 31.5/60, SD 4.3), and 62.6% reported good malaria prevention practice (mean 15.6/27, SD 6.0) (Table 2). At the item level (Additional file 1, Tables S1 to S4), the most consistently practiced measures were sleeping under a mosquito net (71.1% ‘very often’) and closing doors and windows early (70.3%), while indoor insecticide spraying (17.6%), mosquito repellent use (15.0%), and window screening (31.8%) were used far less consistently.

**Table 2.** Level of knowledge, attitude, perception, and malaria prevention practice among participants (N= 620)

| Domain (score range) | Category | n | % | Mean (SD) |
| --- | --- | --- | --- | --- |
| Knowledge (0-10) | Poor (0-5) | 17 | 2.7 | 8.8 (1.3) |
|  | Good (6-10) | 603 | 97.3 |  |
| Attitude (0-21) | Negative ($\leq 10.5$) | 59 | 9.5 | 16.2 (3.7) |
| | Positive ($> 10.5$) | 561 | 90.5 | |
| Perception (0-60) | Negative ($\leq 30$) | 265 | 42.7 | 31.5 (4.3) |
| | Positive ($> 30$) | 355 | 57.3 | |
| Malaria prevention practice (0-27) | Poor (0-13.5) | 232 | 37.4 | 15.6 (6.0) |
|  | Good (13.6-27) | 388 | 62.6 |  |

*SD*, standard deviation. Category cut-points for each domain are defined in Methods (Measures).

### Socio-demographic correlates of knowledge, attitude, perception, and malaria prevention practice

Knowledge, attitude, perception, and practice scores did not differ significantly by sex (Mann-Whitney U, all p > 0.05)(See Additional file 1, Table S5). Education level was significantly associated with attitude (Kruskal-Wallis H(3) = 23.55, p < 0.001), perception (H(3) = 17.02, p= 0.001), and practice (H(3) = 9.28, p = 0.026), but not knowledge (H(3) = 3.80, p = 0.283). The direction of the education effect was non-linear: attitude and perception scores were highest among participants with no formal education and lowest among those with secondary education (Bonferroni-corrected pairwise p-values 0.001 to 0.05), whereas malaria prevention practice was significantly higher among tertiary-educated participants than among those with secondary education (adjusted p = 0.034); other pairwise comparisons for practice were not significant (Table 3).

**Table 3.**
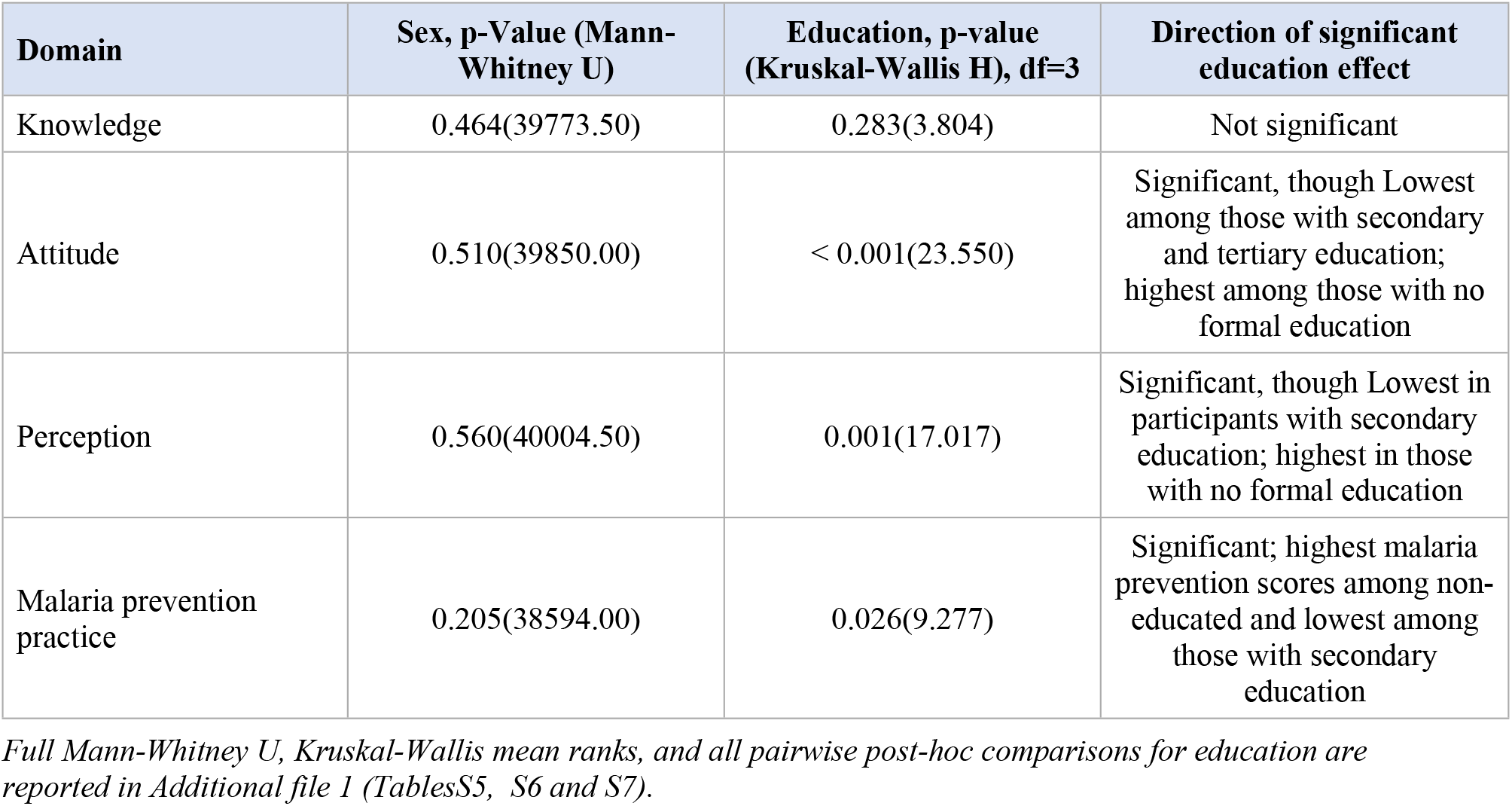
Association of sex and education level with knowledge, attitude, perception, and malaria prevention practice.

### Correlation among knowledge, attitude, perception, and practice

Knowledge was moderately correlated with attitude (rho = 0.514, p < 0.001) and weakly correlated with perception (rho = 0.246, p < 0.001) and malaria prevention practice (rho = 0.158, p < 0.001). Attitude was weakly but significantly correlated with perception (rho = 0.379, p < 0.001) and practice (rho = 0.168, p < 0.001). Perception was not significantly correlated with malaria prevention practice (rho = 0.063, p = 0.115) (Table 4).

**Table 4.** Spearman correlation (rho) among knowledge, attitude, perception, and malaria prevention practice (N = 620)

|  | Attitude | Perception | Malaria prevention practice |
| --- | --- | --- | --- |
| Knowledge | rho = 0.514, p < 0.001 | rho = 0.246, p < 0.001 | rho = 0.158, p < 0.001 |
| Attitude | N/A | rho = 0.379, p < 0.001 | rho = 0.168, p < 0.001 |
| Perception |  | N/A | rho = 0.063, p = 0.115 |

*rho, Spearman's rank correlation coefficient; N/A, not applicable (diagonal/self-comparison).*

### Predictors of malaria prevention practice

In a binary logistic regression model with knowledge, attitude, and perception entered simultaneously, the overall model was significant (chi-square = 37.62, df = 3, p < 0.001). The Hosmer-Lemeshow goodness-of-fit test indicated some deviation from perfect fit (chi-square(8) = 17.995, p = 0.021). Knowledge (AOR = 1.198, 95% CI 1.03-1.40, p = 0.023) and attitude (AOR = 1.101, 95% CI 1.04-1.17, p = 0.001) were independent, significant predictors of good malaria prevention practice: each one-point increase in knowledge score was associated with 19.8% higher odds of good practice, and each one-point increase in attitude score with 10.1% higher odds. Perception was not an independent predictor once knowledge and attitude were accounted for (AOR = 1.00, 95% CI 0.959-1.042, p = 0.994) (Table 5).

**Table 5.** Binary logistic regression of knowledge, attitude, and perception as predictors of good malaria prevention practice.

| Predictor | B (SE) | AOR | 95% CI | p |
| --- | --- | --- | --- | --- |
| Knowledge | 0.181 (0.080) | 1.198 | 1.025-1.400 | 0.023 |
| Attitude | 0.096 (0.029) | 1.101 | 1.040-1.165 | 0.001 |
| Perception | 0.000 (0.021) | 1.000 | 0.959-1.042 | 0.994 |
| Constant | -2.603 (0.790) | 0.074 | N/A | 0.001 |

*AOR, adjusted odds ratio; SE, standard error; CI, confidence interval; N/A, not applicable (no confidence interval is reported for the model constant).*

## Discussion

This study demonstrated that the scourge of HIV infection predominantly affects young people and females, and this same trend is reflected in national and global statistics, including ART clinics in other countries(3–5,21–23). In this HIV-positive population, knowledge and attitude, but not perception, independently predicted malaria prevention practice, and practice remained suboptimal despite very high knowledge and largely positive attitudes. This pattern is broadly consistent with studies in other HIV-positive populations that found knowledge of net use predicted uptake [10] and that positive attitude promoted malaria prevention in the general population [12], but it extends that literature by showing, for the first time in an HIV-positive cohort, that knowledge and attitude retain independent predictive value even after accounting for each other and for perception.

Perception was positive in only 57.3% of participants and, unlike knowledge and attitude, did not independently predict practice. This does not mean perception is irrelevant, but other environmental and socio-cultural factors may be responsible. Additionally, perception was moderately correlated with attitude (rho = 0.379) and knowledge (rho = 0.246), so some of its influence may already be captured by attitude and Knowledge in the adjusted model. It is also consistent with findings that perception has been comparatively underexplored and inconsistently associated with practice in general-population KAP studies [13,14]. Sleeping under mosquito nets and closing windows were commonly used methods in this study, similar to studies in other ART clinics(22) though attributed to their availability following free distribution by their government(22,30,31). Perceived barriers were common in this cohort, including fear of side effects of insecticides impregnated in nets or chemoprevention with repellents, residual spraying, and inconvenience of clinic attendance, which suggests that structural or health-system barriers, such as consistent supply of nets and access to spraying or repellents, and affordability of newer prevention methods like repellents, may matter more than individual perception once knowledge and attitude are accounted for. These barriers were also observed in a study in which the newer prevention methods were not embraced due to side effects, lack of knowledge, and affordability(32).

Contrary to the common assumption that more schooling produces more favourable knowledge, attitude, and perception profiles [12], attitude and perception mean rank scores towards malaria prevention practices, in this cohort, were highest among participants with no formal education and lowest among those with secondary education, while practice was highest among none-educated participants. This pattern should be interpreted cautiously: the no-education subgroup was small (n = 25), and Likert-scale attitude and perception items may have been interpreted differently across education levels. It nonetheless suggests that educational attainment alone does not straightforwardly predict favourable knowledge, attitude, and perception profiles about malaria prevention in this population, and that assumptions built into education-stratified health messaging should be tested rather than presumed, as observed in the Know-do approaches(33). Additionally, malaria prevention information is spread on media and in an early morning educational program at the ART clinic every day before the start of the day without regard for education levels or demographic characteristics, as observed during data collection at the site, and this may explain the observed results.

This study has several limitations. Self-reported practice is subject to social desirability and recall bias, which trained, neutral interviewers were used to mitigate but could not eliminate. The single-centre, urban design limits generalisability to other HIV care settings, particularly rural clinics; this is a genuine limitation given that comparable HIV-specific data are lacking, so readers should not extrapolate beyond similar urban ART populations. Knowledge, attitude, perception, and practice scores were dichotomised at fixed scale midpoints rather than at empirically derived or previously validated cut-points, which may have reduced statistical power or exaggerated apparent differences between groups. Perception was retained in the adjusted regression model despite a non-significant bivariate correlation with practice because all three variables are core to the KAP and Health Belief Model constructs; its null adjusted result should be read as adding no independent explanatory value beyond knowledge and attitude in this sample, not as evidence that perception plays no role in malaria prevention generally. Finally, the cross-sectional design precludes causal inference.

## Conclusions

Despite near-universal knowledge (97.3%) and predominantly positive attitudes (90.5%) toward malaria prevention, only 62.6% of HIV-positive participants in this urban ART clinic reported good malaria prevention practice, and only knowledge and attitude, not perception, independently predicted malaria prevention practice. This gap between what participants knew and felt and what they actually did suggests that HIV care programmes seeking to reduce malaria burden in this population should pair knowledge-based health education with structural support for the tools and access needed to act on that knowledge, rather than relying on education alone.

## Data Availability

The dataset(s) supporting the conclusions of this article are available from the corresponding author on reasonable request.

## Declarations

### Ethics approval and consent to participate

The study was conducted in accordance with the Declaration of Helsinki and adhered to applicable ethical guidelines and regulations for research involving human participants in Uganda. Ethical approval was obtained from the Research Ethics Committee at Kabale University (protocol KABREC-2025-654), with additional clearance from the research committee of China-Uganda Friendship Hospital, Naguru. Written informed consent was obtained from all participants; assent and parental or guardian consent were obtained for participants under 18 years.

### Human Ethics and consent to participate

The study was approved by the relevant institutional review board of Kabale University, and consent or assent with parental or guardian consent was obtained.

### Consent for publication

Not applicable. The manuscript contains no person’s data, images, or case details.

### Clinical Trial number

Not Applicable

### Authors’ contributions

AA conceived and designed the study, collected and analysed the data, wrote the original research paper, provided study resources, and drafted the first draft of the manuscript. KM supervised and provided oversight to the overall research and contributed to study design. MJ provided expert knowledge, made tables in additional file 1, and contributed to the overall manuscript write-up. All authors read and approved the final manuscript.

## Acknowledgements

On behalf of all co-authors, we acknowledge the support of Juliet Bandaru as the first supervisor during the inception of the research and Tuhairwe Richard for proofreading and providing expert opinion during the research.

Appreciation to the staff at the ART Clinic of China Uganda Friendship Hospital for their support during data collection.

